# COMPARATIVE IMMUNOGENICITY OF BNT162b2 mRNA VACCINE WITH NATURAL COVID-19 INFECTION

**DOI:** 10.1101/2021.06.15.21258669

**Authors:** Mina Psichogiou, Andreas Karabinis, Garyphallia Poulakou, Anastasia Antoniadou, Anastasia Kotanidou, Dimitrios Degiannis, Ioanna D. Pavlopoulou, Antigoni Chaidaroglou, Sotirios Roussos, Elpida Mastrogianni, Irene Eliadi, Dimitrios Basoulis, Konstantinos Petsios, Konstantinos Leontis, Eleni Kakkalou, Konstantinos Protopapas, Edison Jahaj, Maria Pratikaki, Konstantinos N. Syrigos, Pagona Lagiou, Helen Gogas, Sotirios Tsiodras, Gkikas Magiorkinis, Dimitrios Paraskevis, Vana Sypsa, Angelos Hatzakis

## Abstract

The mRNA vaccine BNT162b2 has proven highly effective and currently many millions are being vaccinated. There are limited and conflicting data from immunogenicity studies on the effects of age, gender, vaccination side effects (VSE), risk factors for severe COVID-19 (RFS-COV), obesity (BMI) and previous SARS-CoV-2 (Pr-CoV) Moreover, immunogenicity data from COVID-19 patients comparing various disease categories of natural infection i.e. asymptomatic vs mild vs moderate vs severe infection are sparse, and include limited number of individuals.

This study included 871 vaccinated health care workers (HCW) and 181 patients with natural infection. Immunogenicity was assessed by a quantative assay measuring anti-SARS-CoV-2 against the RBD domain of the spike protein (anti-RBD) and anti-SARS-CoV-2 against nucleocapsid protein (anti-N). Samples were collected 1-2 weeks after completion of the 2^nd^ dose in the vaccinated HCWs and 15-59 days post symptoms onset in patients with natural infection.

The concentration of anti-RBD in vaccinated individuals after multivariable analysis was significantly associated with age, gender, VSE and Pr-CoV. Specifically, anti-RBD median levels (95% CI) were lower by 2,466 (651-5,583), 6,228 (3,254-9,203) and 7,651 (4,479-10,823) AU/ml in 35-44, 45-54, 55-70 yrs respectively, compared with 18-34 yrs group. In females, median levels of anti-RBD were higher by 2,823 (859-4,787) compared with males, in individuals with VSE were higher by 5,024 (3,122-6,926) compared with no VSE, and in HCWs with Pr-CoV were higher by 9,971 (5,158-14,783) AU/ml compared with HCWs without Pr-CoV.

Among individuals with natural infection, the median anti-RBD levels were 14.8 times higher in patients with critical COVID-19 infection compared with non-hospitalized individuals. The ratio of anti-RBD in vaccinated individuals versus those with natural infection varied from 1.0 up to 19.4 according to the clinical subgroup of natural infection.

This study proves the high immunogenicity of BNT162b2 vaccine although its sustainability remains to be seen. The use of comparative data from natural infection serological panels, expressing the clinical heterogeneity of natural infection may facilitate early decisions for vaccine evaluation in clinical trials.

## INTRODUCTION

Coronavirus disease 2019 (COVID-19) is expanding despite mitigation policies of variable success. By the end of 2020, an increasing number of safe and effective vaccines were approved, and large vaccination programs are underway around the world. By May 15, 2021 more than 162,500,000 cases and 3,371,000 deaths were reported while more than 1.41 billion vaccine doses were administered.

The first approved vaccine was Pfizer - BNT162b2, a lipid nanoparticle – formulated mRNA vaccine encoding the SARS-CoV-2 full length spike modified by two proline mutations. Preliminary findings among healthy men and women showed that two 30mg doses elicited high SARS-CoV-2 neutralizing antibody titers and robust antigen-specific CD8+ and Th1-type CD4+ T-cell responses **[1]**. Moreover, in a multinational, placebo -controlled observer blinded efficacy trial including 43.548 healthy or with stable chronic conditions participants, the vaccine was found safe with an efficacy of 95% (95% credible interval 90.3-97.6%) in preventing symptomatic COVID-19 disease ≥7 days after the 2nd dose **[2]**. Similar vaccine efficacy (90-100%) was observed across groups defined by age, sex, race, baseline body mass index (BMI) and the presence of coexisting conditions. History of previous COVID-19 treatment, immunosuppressive therapy or diagnosis with an immunocompromising condition were exclusion criteria in this pivotal study.

A second approved mRNA vaccine, Moderna – mRNA 1273, showed similar efficacy 94.1% (95% 89.3 – 96.8) in preventing COVID-19 illness **[3]**.

The BNT162b2 was also evaluated in a mass vaccination campaign in Israel with an efficacy, 7 days from the 2nd dose, of 94%, 87% and 92% in preventing symptomatic disease, hospitalization, and severe disease, respectively **[4]**.

Presence of neutralizing antibodies is a strong correlate of vaccine efficacy, although a protection threshold is not established. However, measuring neutralizing antibodies in a large scale is challenging. The development of binding assays directed against spike protein of SARS-CoV-2 showed excellent correlation with neutralizing antibodies **[5-10]** and it, gives the opportunity to assess the immunogenicity over time of SARS-CoV-2 vaccines in large scale. Assessment of vaccine immunogenicity to predict and to monitor vaccine effectiveness is important in groups of individuals not included in clinical trials such as patients with immunocompromising conditions **[11]**.

Despite the high levels of vaccine efficacy in immunocompetent individuals the duration of BNT162b2 protection remains unknown. Antibody titers in other coronaviruses (seasonal, SARS CoV-1, MERS) wane over time and this is the case with COVID-19 antibodies in natural infection **[12-14]**.

Natural infection protects from reinfection for at least 7 months, and it represents a benchmark for comparison with vaccine efficacy and immunogenicity **[15-17]**. Higher levels of NA and binding antibodies have been associated with increased clinical severity of natural infection in several studies **[13,14,18,19]**.

Phase I/II SARS-CoV-2 vaccine immunogenicity studies on approved or under approval vaccines included limited numbers of individuals with natural infection as a control group **[1, 20-23]**. However, the heterogeneity of natural infection was not considered and comparisons of vaccinated individuals with the former group were incomplete.

Herein, we report comparative immunogenicity data of BNT162b2 mRNA vaccine after the 2^nd^ dose with a large cohort of individuals with natural COVID-19 infection.

## METHODS

### Vaccination for HCWs

Participants were vaccinated with 2 doses of BNT162b2 21 days apart. The vaccine was administered intramuscularly and included 30 mg of SARS-CoV-2 full length lipid nanoparticle formulated mRNA.

The study was designed to assess immunogenicity at time intervals 1-2 weeks after the 2^nd^ dose (28-35 days) and 4, 6, 8, and 12 months after the 1^st^ dose. Immunogenicity 1-2 weeks after the 2^nd^ dose was expected to be highest, based on the results from phase I/II studies **[1]**. HCWs from 2 teaching hospitals, Laiko General Hospital (Hospital 1) and Onassis Cardiac Surgery Center (Hospital 2) were informed about the study and participated after signing an informed consent. A brief questionnaire was administered to HCWs with information about age, gender, education, position within hospital, BMI, history of risk factors for severe COVID-19 (RFS-CoV), previous COVID-19 (Pr-CoV) and history of adverse reactions after vaccination (VSE).

### Natural Infection Group

A group of 315 patients with natural SARS-CoV-2 infection diagnosed with RT-PCR testing was included in the study. Participants provided informed consent. Age, gender, diagnostic tests, symptoms, hospitalization, disease severity **[24]** and admission in intensive care unit (ICU) were recorded. The present analysis includes in total 180 patients, 157 hospitalized and 23 non-hospitalized, 171 symptomatic and 9 asymptomatic individuals, 155 patients with available time from symptom onset (PSO) and 163 with estimation of severity.

The study was approved by the IRB committees of Laiko General Hospital and Onasis Cardiac Surgery Center.

### Serological Tests

Serum samples collected after venipuncture, were tested for SARS-CoV-2 IgG binding antibodies to nucleocapsid protein (anti-N) and anti-SARS-CoV-2 receptor binding domain (RBD) spike protein IgG (anti-RBD).

The first assay is a qualitative one with an index [sample/calibrator (s/c)] cutoff of 1.4. Samples with an index ≥1.4 are considered positive and <1.4 negative. The clinical sensitivity of the anti-N assay in samples collected ≥15 days after onset of symptoms is 100% (95% CI 95.9-100%) and the clinical specificity 99.63% (95% CI 99.05-99.90%), according to the manufacturer **[25]**.

The second assay (Abbott SARS-CoV-2 IgG II Quant) or anti-RBD was used to quantify IgG antibodies against the receptor binding domain (RBD) of the S1 subunit of the spike protein. The linear range is between 21 and 40,000 AU/ml. The lower limit of detection was 6.8 AU/ml and the reportable interval 6.8-80,000 AU/ml. The clinical sensitivity was 98.81% (95% CI 93.56-99.94%) in samples collected ≥15 days after the positive PCR and the clinical specificity 99.55% (95% CI 99.15-99.76%), at a cutoff value 50 AU/ml **[25]**.

Both assays are based on chemiluminescent microparticle immune assay (CLIA).

The correlation coefficient in weighted linear regression of WHO standard with the Abbott anti-RBD is 0.999, and transformation of Abbott anti-RBD AU/ml to WHO BAU/ml is feasible using the equation BAU/ml = 0.142 x AU/ml **[25]**.

## STATISTICAL ANALYSIS

Median values, 25th and 75th percentiles were used to describe anti-RBD levels. We compared levels between vaccinated health care workers and individuals with a prior SARS-CoV-2 infection diagnosis or other groups using the non-parametric Mann–Whitney U test or Kruskal Wallis test. Multiple linear regression was used to identify factors associated with anti-RBD levels. Ratios of the median anti-RBD levels among vaccinated people versus the median levels among persons with natural immunity (asymptomatic, mild symptoms and moderate/severe symptoms) were also calculated.

We conducted all statistical analyses using STATA 13.1. Figures were created in R (v 4.1.0).

## RESULTS

### Vaccinated HCWs

Eight hundred seventy-one HCWs participated in the study. Their sociodemographic characteristics are shown in **Table 1a**.

**Table 1.**
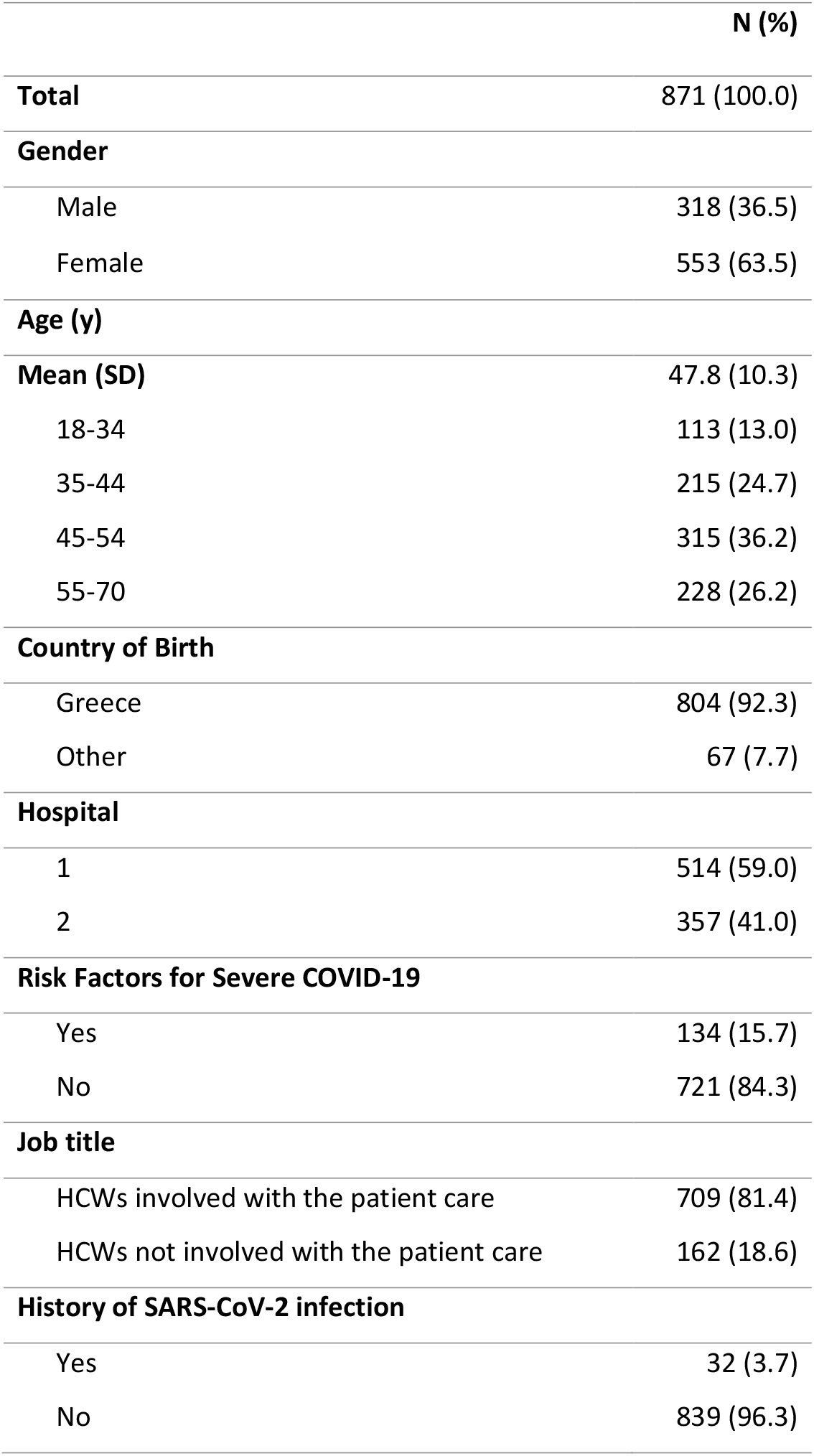
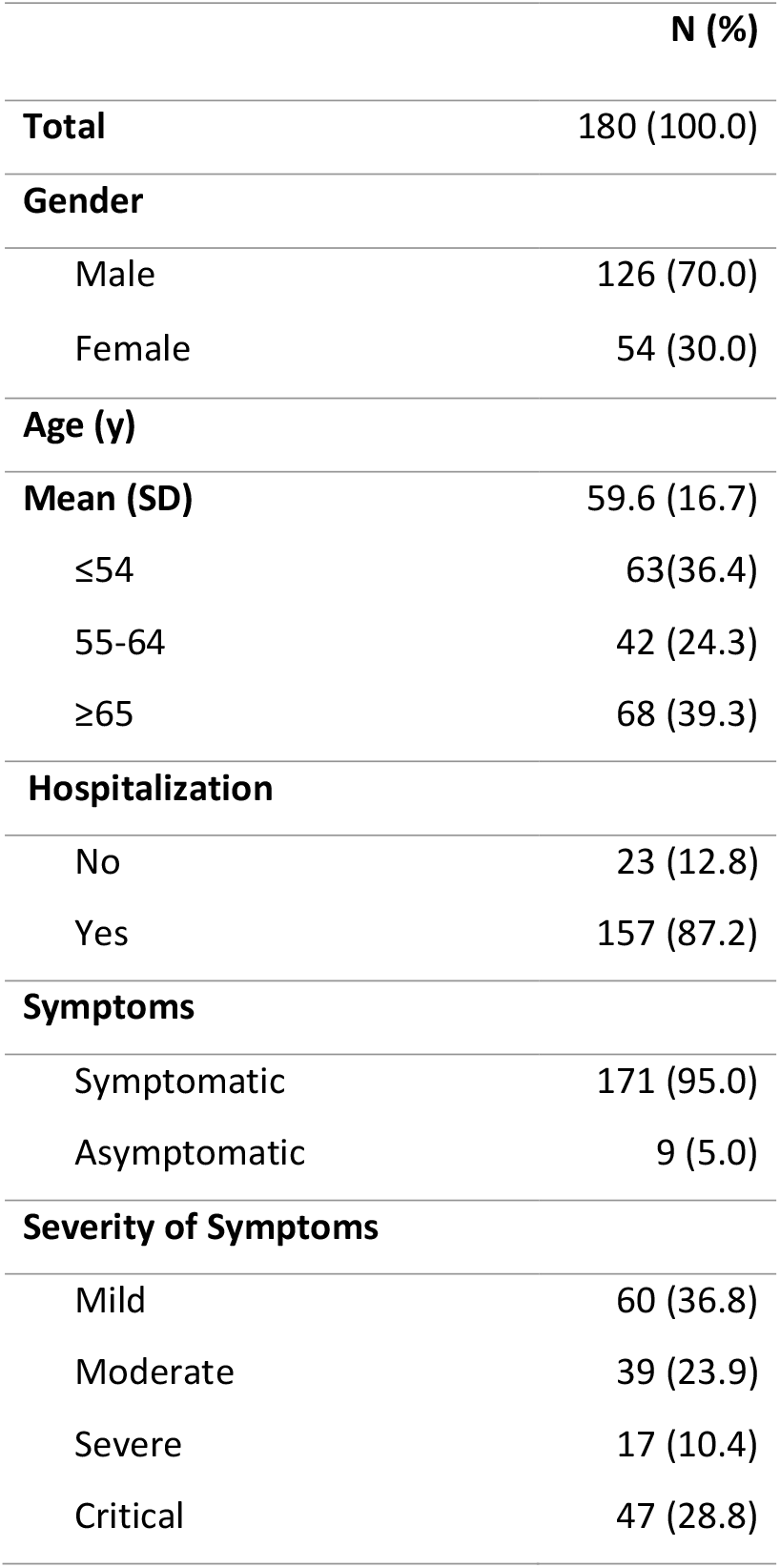
Sociodemographic and clinical characteristics of participants in immunogenicity studies. Table 1a: Health care workers. Table 1b: Individuals with COVID-19 infection

The prevalence of the SARS-CoV-2 anti-N IgG was 3.7% (32 out of 871) (95% CI 2.5-5.2%) and of the anti-RBD IgG was 99.7% (868 out of 871) (95% CI 99.0-99.3%). The concentrations of anti-RBD ranged from <6.8 up to higher than 80.000 AU/ml. The 2.5^th^, 50^th^ and 97.5^th^ percentiles of anti-RBD were 1,680, 15,877 and 55,309 AU/ml, respectively **(Supp. Figure 1)**. The median (IQR) anti-RBD levels by age, gender, country of birth, BMI, risk factors for severe COVID-19, side effects of vaccination and previous SARS-CoV-2 infection are shown in **Table 2**. Gender, age, previous SARS-CoV-2 infection, side effects of vaccination and risk factors for COVID-19 showed statistically significant association with concentrations of anti-RBD. However, in a multivariable linear regression analysis only gender, age, side effects of vaccination and previous SARS-CoV-2 infection, showed statistically significant associations with concentrations of anti-RBD **(Table 2)**. More specifically, females had on average 2,823 (95% CI 859-4,787) AU/ml concentration higher than males HCWs (p = 0.05). Participants aged 55-70 yrs had on average 7,651 (95% CI 4,479-10,823) AU/ml lower than HCWs 1834 yrs (p<0.001).

**Table 2.**
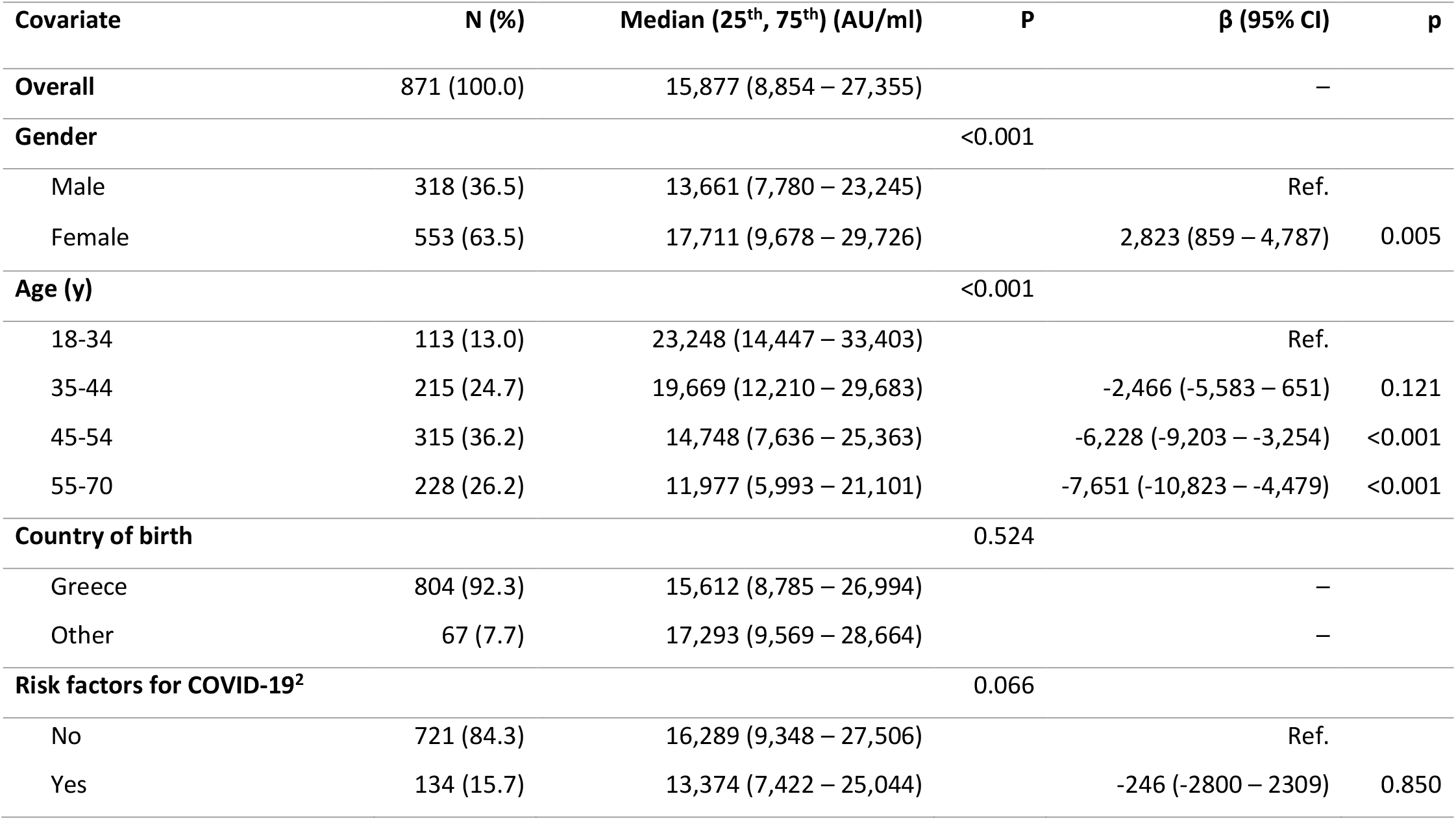
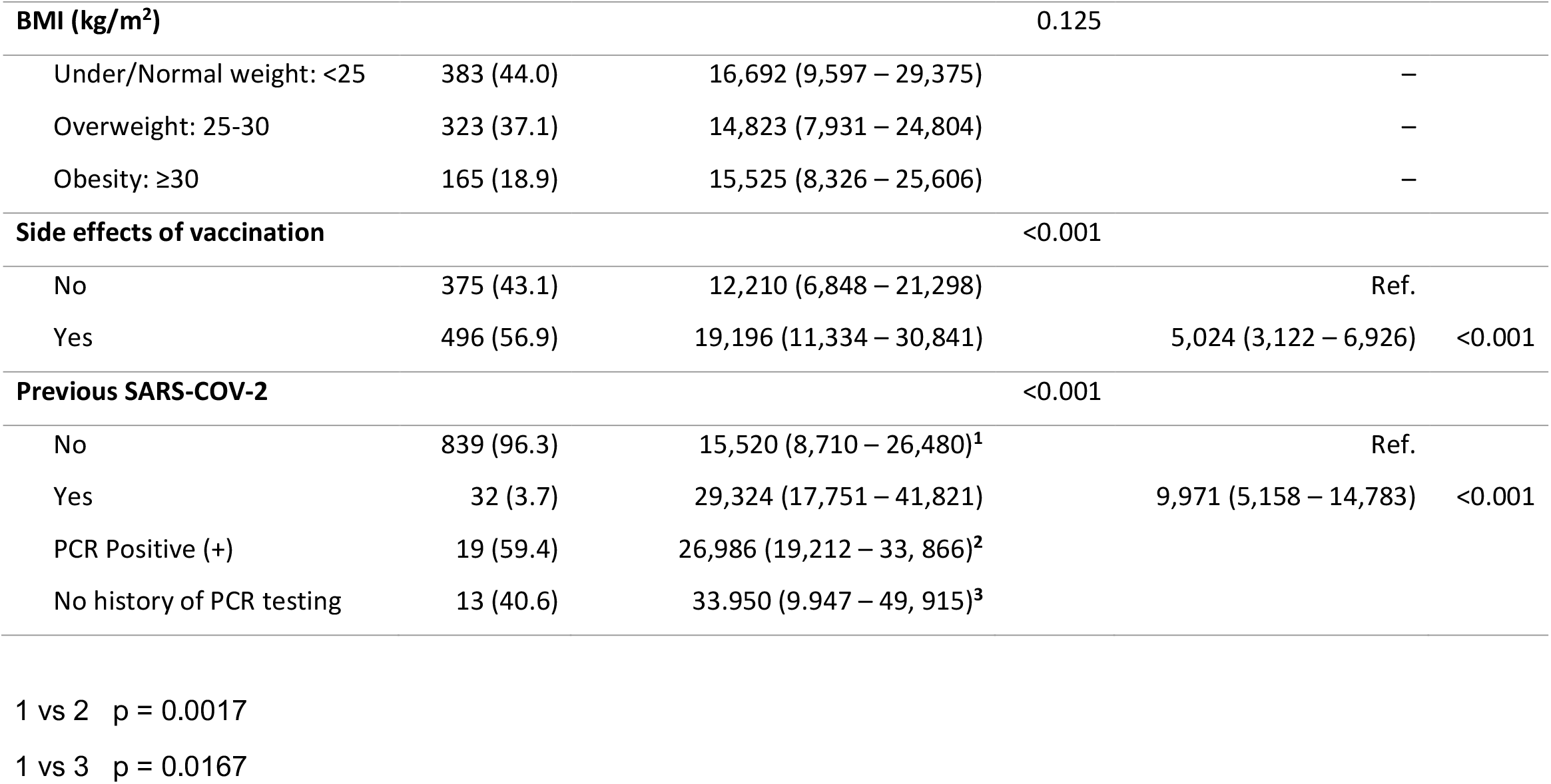
Median (25^th^, 75^th^) concentration of anti-SARS-CoV-2 IgG-II antibodies after the second dose of BNT162b2 vaccine and coefficients (β) along with 95% Confidence Intervals from multiple linear regression.

**Figure 1:**
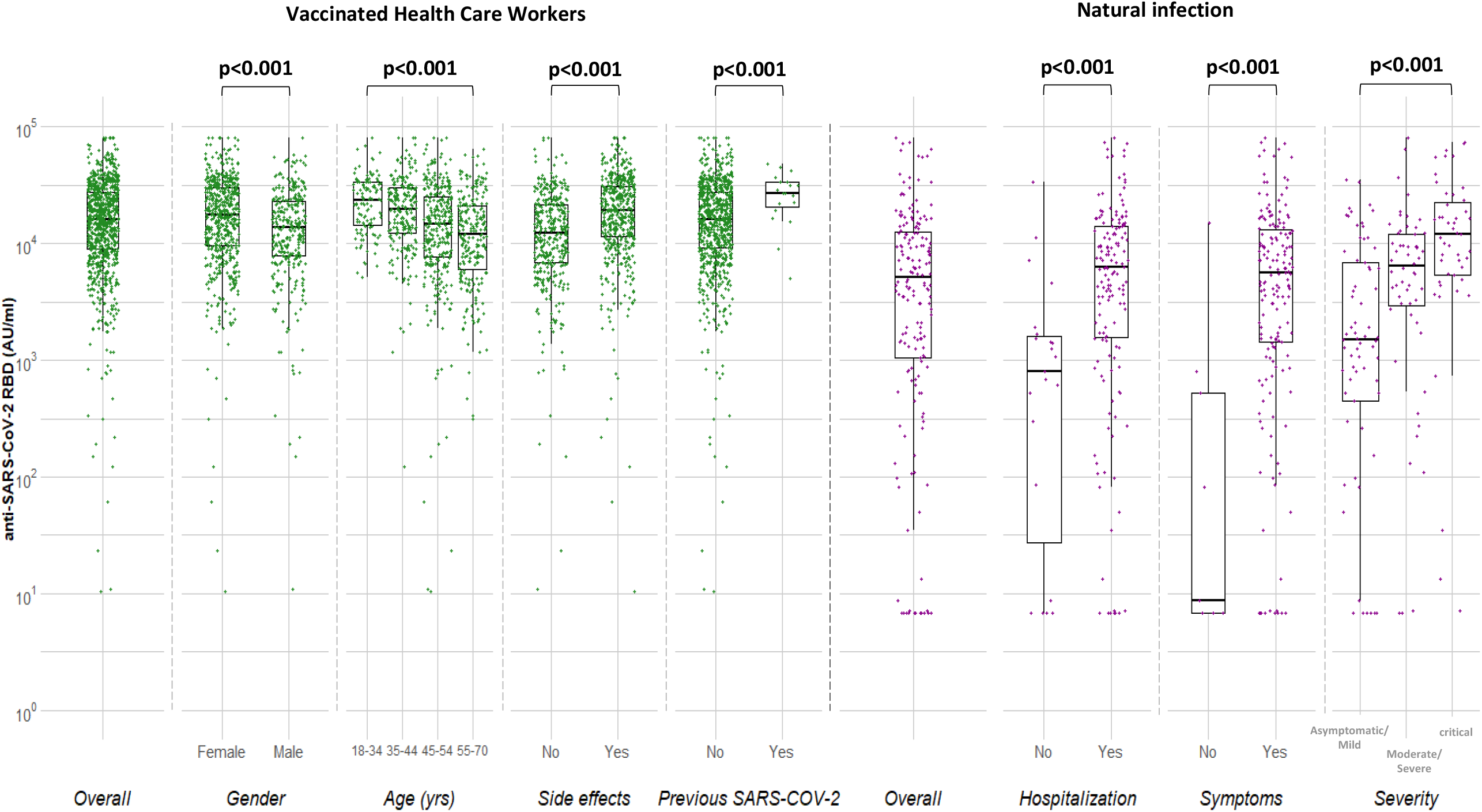
Median concentrations of anti-SARS-CoV-2 RBD (AU/ml) in vaccinated health care workers 7-15 days after the 2nd dose of BNT162b2 and individuals with natural infection.

HCWs reporting side effects had a concentration of 5,024 (95% CI 3,122-6,926) AU/ml higher than those non-reporting side effects (p<0.001). Moreover, HCWs with previous SARS-CoV-2 had higher levels by 9,971 (95% 5,158-14,783) AU/ml compared to COVID-19 naïve individuals (p<0.001).

Time from 2^nd^ dose: The median levels of anti-RBD were calculated according to the time from the 2^nd^ dose. The maximum levels were reached 11 days after the 2^nd^ dose while a sharp reduction was observed 15-17 days after the 2^nd^ dose (Kruskal-Wallis, p=0.007) **(Suppl. Table 1)**. Multivariable analysis showed that reduction was independent of age, gender, side effects of vaccination and previous SARS-CoV-2 infection (data not shown).

### Natural Infection

The early convalescent samples post-symptoms onset (PSO) ≥15-59 days of symptomatic (n=155), asymptomatic individuals (n=9), hospitalized (n=157) and non-hospitalized (n=23) individuals were included in this analysis. The sociodemographic and clinical characteristics are shown in **Table 1b**.

The prevalence (95% CI) of anti-N and anti-RBD was 88.3% (82.7-92.6%) and 90.6% (85.3-94.4%) respectively.

The median (IQR) anti-RBD levels by age, gender, symptomatic/asymptomatic, severity of clinical disease and time POS are shown in **Table 3**. The median (IQR) anti-RBD concentrations were 9 (<6.8-520) and 5,547 (1,415-13,325) AU/ml in asymptomatic and symptomatic individuals respectively (p<0.001), 6,271 (1,583-14,121) and 808 (9-1,668) hospitalized and non-hospitalized respectively.

**Table 3.**
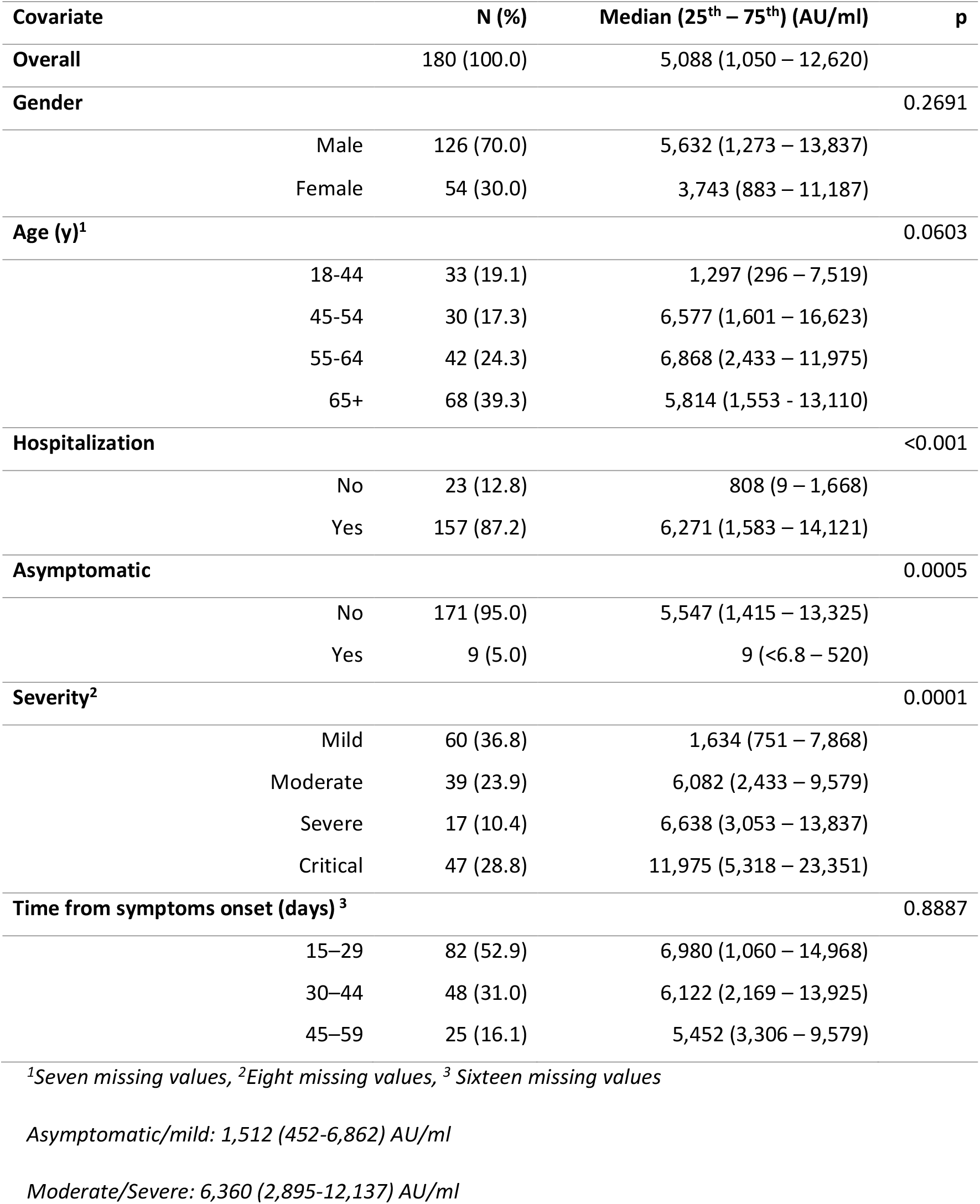
Median (25th, 75th) concentration of anti-SARS-CoV-2 IgG-II antibodies in symptomatic SARS-CoV-2, 15-59 days after infection and asymptomatic individuals.

| Covariate |  | N (%) | Median (25<sup>th</sup> – 75<sup>th</sup>) (AU/ml) | p |
| --- | --- | --- | --- | --- |
| **Overall** |  | 180 (100.0) | 5,088 (1,050 – 12,620) |  |
| **Gender** |  |  |  | 0.2691 |
|  | Male | 126 (70.0) | 5,632 (1,273 – 13,837) |  |
|  | Female | 54 (30.0) | 3,743 (883 – 11,187) |  |
| **Age (y)<sup>1</sup>** |  |  |  | 0.0603 |
|  | 18-44 | 33 (19.1) | 1,297 (296 – 7,519) |  |
|  | 45-54 | 30 (17.3) | 6,577 (1,601 – 16,623) |  |
|  | 55-64 | 42 (24.3) | 6,868 (2,433 – 11,975) |  |
|  | 65+ | 68 (39.3) | 5,814 (1,553 – 13,110) |  |
| **Hospitalization** |  |  |  | <0.001 |
|  | No | 23 (12.8) | 808 (9 – 1,668) |  |
|  | Yes | 157 (87.2) | 6,271 (1,583 – 14,121) |  |
| **Asymptomatic** |  |  |  | 0.0005 |
|  | No | 171 (95.0) | 5,547 (1,415 – 13,325) |  |
|  | Yes | 9 (5.0) | 9 (<6.8 – 520) |  |
| **Severity<sup>2</sup>** |  |  |  | 0.0001 |
|  | Mild | 60 (36.8) | 1,634 (751 – 7,868) |  |
|  | Moderate | 39 (23.9) | 6,082 (2,433 – 9,579) |  |
|  | Severe | 17 (10.4) | 6,638 (3,053 – 13,837) |  |
|  | Critical | 47 (28.8) | 11,975 (5,318 – 23,351) |  |
| **Time from symptoms onset (days)<sup>3</sup>** |  |  |  | 0.8887 |
|  | 15–29 | 82 (52.9) | 6,980 (1,060 – 14,968) |  |
|  | 30–44 | 48 (31.0) | 6,122 (2,169 – 13,925) |  |
|  | 45–59 | 25 (16.1) | 5,452 (3,306 – 9,579) |  |

<sup>1</sup>Seven missing values, <sup>2</sup>Eight missing values, <sup>3</sup> Sixteen missing values
*Asymptomatic/mild: 1,512 (452-6,862) AU/ml*
*Moderate/Severe: 6,360 (2,895-12,137) AU/ml*

The median (IQR) anti-RBD levels were highly associated with increased severity; <u>Mild</u>: 1,634 (751-7,868), <u>Moderate</u>: 6,082 (2,433-12,224), <u>Severe</u>: 6,638 (3,053-13,837) and

<u>Critical</u> 11,975 (5,138-23,351) AU/ml (p<0.001 for between group comparisons). Hospitalized individuals had 7.8-fold times higher median anti-RBD levels than non-hospitalized. Specifically, patients with moderate, severe and critical disease had 4.0, 4.4, 7.9-fold times higher anti-RBD level, than those with asymptomatic/mild infection, respectively **(Figure 2)**.

**Figure 2:**
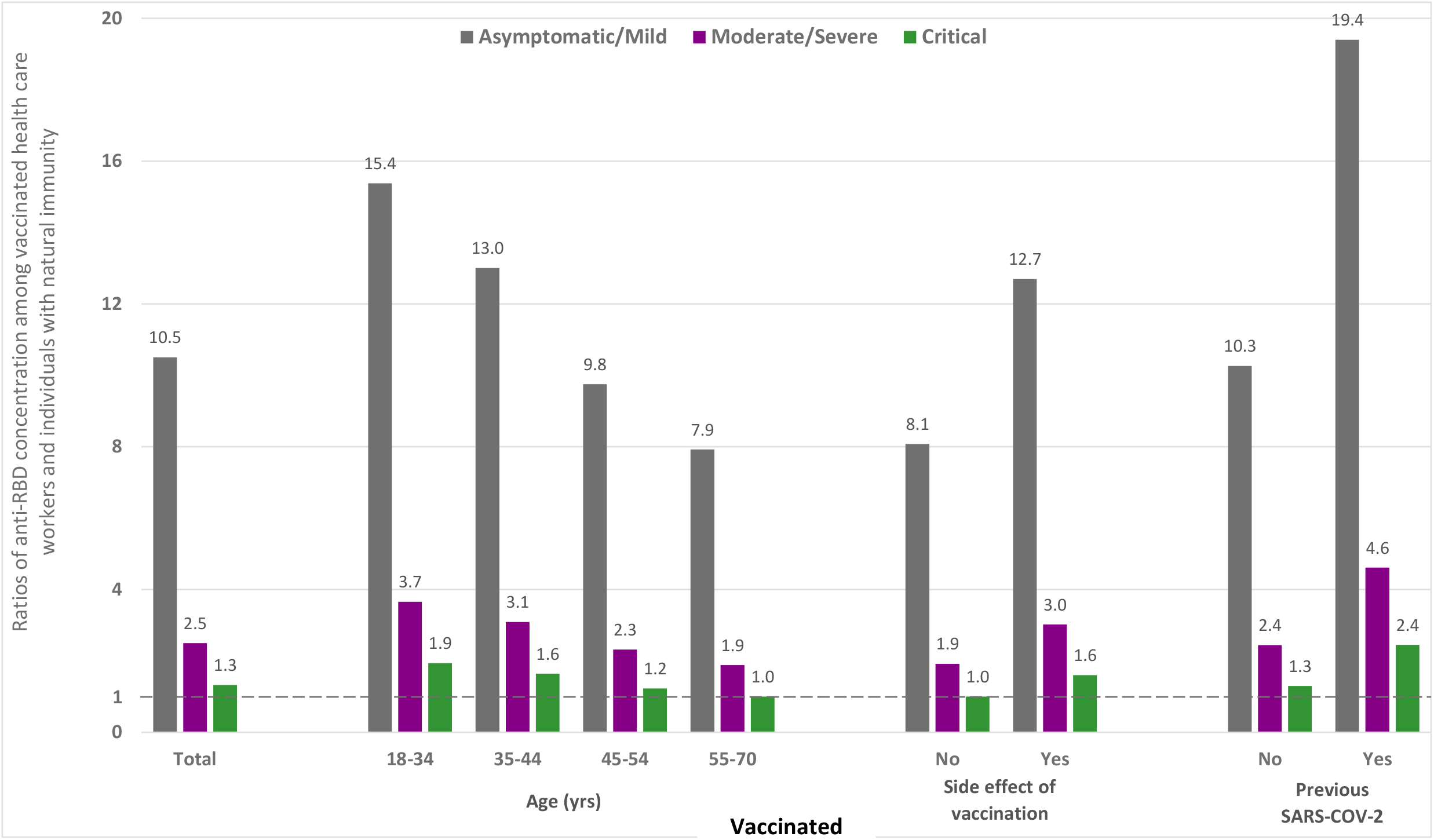
Ratio of median concentrations of anti-SARS-CoV-2 RBD in vaccinated groups versus naturally infected individuals with asymptomatic/mild, moderate/severe and critical infection.

### Comparison of anti-RBD levels in vaccinated HCWs and in individuals with natural infection

The ratio of median anti-RBD levels in vaccinated after the 2^nd^ dose versus the median levels of those with natural infection in the early convalescent period 15-59 days POS is shown in **Figure 2**. Anti-RBD concentrations of natural infection were used as denominators (asymptomatic/mild, moderate/severe and critical infection).

We observed several fold differences in the anti-RBD ratio for each vaccinated group e.g., across different age groups, i.e., 18-34, 35-44, 45-54, 55-70 years old, and the ratio of median anti-RBD levels for vaccinated over natural infection was 1.9-15.4, 1.6-13.0, 1.2-9.8 and 1.0-7.9, respectively. In the group with VSE the ratio was 1.6-12.7 and in the group with Pr-CoV 2.4-19.4 **(Figure 2)**. For the whole group of vaccinated individuals, the ratio was 1.3, 2.5, 10.5 based on critical, moderate/severe and asymptomatic/mild patients with natural infection respectively.

## DISCUSSION

Several lines of evidence suggest that neutralizing antibodies are correlates of protection (CoP) against SARS-CoV-2 infection:

a. Studies in macaques infected by SARS-CoV-2 demonstrated the pivotal role of NA and S specific CD4^+^ and CD8^+^ T-Cell responses to provide near complete protection in rechallenge experiments **[26,27]**.
b. Studies in non-human primates vaccinated by SARS-CoV-2 vaccines demonstrated an NA threshold for complete protection **[28-30]**.
c. A study of natural infection outbreak in a fishery vessel where of the 117 individuals who were seronegative to NA and binding antibodies prior to departure, 104 were (88.9%) infected by RT-PCR while from the 3 crew members with presence of NA, anti-RBD and antibodies to full-length spike, none was infected **[31]**.
d. Prospective study in 3.168 marine recruits revealed aggregate infection of 48% during a 6-week training. Among 189 anti-spike and anti-RBD positive 19 (10%) were infected by RT-PCR during training. Lower levels of neutralizing and binding antibodies were associated with higher incidence of infection **[32]**.
e. A higher AZD1222 vaccine efficacy was demonstrated with higher levels of NA and anti-spike IgG in a vaccination trial **[33]**. An excellent correlation of anti-RBD and NA was observed in vaccination trials **[33-37]** or other studies **[6,8-10, 38]**.

A remarkable finding of mRNA vaccines immunogenicity studies is that levels of anti-RBD and neutralizing antibodies titers do not change after the 2^nd^ dose in individuals previously infected, suggesting that the 2^nd^ dose of BNT122b2 or other vaccines may not be necessary in previously infected immunocompetent individuals **[39-40]**. In our study we tested vaccinated HCWs by anti-N and anti-RBD 1-2 weeks after the 2^nd^ dose. Thirty-two HCWs were found positive for anti-N for a prevalence of 3.2% (32/871) (95% CI 2.5 – 5.2%). Only 19/32 (59.4%) had a history of previous COVID-19 diagnosed by RT-PCR suggesting that anti-N is a useful test to assess previous COVID-19 infection during vaccination. Both groups of anti-N positive subjects had significantly higher anti-RBD levels compared with HCWs without previous SARS-CoV-2 infection.

Studies of BNT162b2 immunogenicity after 1^st^ and 2^nd^ dose found various associations with age, gender, obesity, vaccination side effects and previous COVID-19. However, confounding effects were not controlled by multivariable analysis **[41-46]**. In our study anti-RBD levels significantly decreased with older age, male gender, presence of risk-factors for COVID-19 and increased with side-effects and previous SARS-CoV-2. After multivariable analysis, significant association remained with age, gender, side-effects of vaccination and previous COVID-19.

We further studied a large group of individuals with natural infection including 175 symptomatic and 26 asymptomatic diagnosed with RT-PCR and available demographic and clinical information. Sera were collected 15-59 days POS in symptomatic individuals. To our knowledge, this is the largest and more comprehensive natural infection panel included in SARS-CoV-2 vaccine immunogenicity studies. The immunogenicity difference of moderate/severe and asymptomatic infection exceeds 1 log10 and it may bias immunogenicity comparisons in vaccine studies. By using asymptomatic infection as baseline group, the anti-RBD concentration was several folds higher in vaccinated individuals compared with natural infection. On the contrary by using moderate/severe infection the vaccinated individuals had slightly elevated concentrations of anti-RBD compared to patients with natural infection.

Overall, these data document the high immunogenicity of BNT162b2 vaccine in comparison with natural infection. Both mRNA BNT1162b2 and mRNA 1273 vaccines are highly immunogenic although this is not the case with several vaccines under evaluation. In a vaccine study using detailed clinical information in the natural infection group, the ratio of anti-RBD and NA in vaccinated vs natural infection was below 1 **[47]**.

Emergence of novel SARS-CoV-2 variants harboring mutations that can either enhance transmissibility or reduce neutralization activity of vaccine sera are of major concern and include the B.1.1.7, B.1.351, P.1 and B.1.617.2 documented first in the UK, South Africa, Brazil and India, respectively. The effectiveness of the BNT162b2 vaccine was assessed under real-world conditions in Qatar, where B.1.1.7 and B.1.351 variants were co-circulating **[48]**. Specifically, the effectiveness of the vaccine against the B.1.1.7 and B.1.351 documented infections was 89.5% (95% CI, 85.9 to 92.3) and 75.0% (95% CI, 70.5 to 78.9), respectively, thus suggesting that mRNA vaccine can protect even against immune escape variants of concerns, such as the B.1.351.

The present analysis assumes according to findings from previous reports that NA and anti-RBD are correlates of protection against SARS-CoV-2. Although the mechanism of protection is complex including several aspects of humoral and cell-mediated immunity, the role of neutralizing antibodies seems profound. The use of multiple NA assays, many of them requiring increased biosafety laboratories is a barrier for large clinical studies. The use of anti-RBD or other antibodies against spike protein may accelerate the study of pathogenesis of SARS-CoV-2 and provide useful tools for assessing vaccine efficacy and their effectiveness over time. Moreover, standardized sera panel with natural infection may be proven valuable for predicting vaccine efficacy without conducting expensive and time-consuming randomized phase 3 clinical trials **[49]**.

This study has some strengths including the large number of vaccinated individuals evaluated, the use of a large natural infection panel and the use of a validated, commercially available assay for anti-RBD testing. The major limitation is the lack of cell-mediated immunity tests to capture aspects of T and B cell immunity.

Prospective evaluation of vaccine immunogenicity in large, vaccinated cohorts is underway and the comparison, with prospective data from natural infection may clarify important questions of COVID-19 pathogenesis and vaccine effectiveness over time.

## Data Availability

A limited data set is available for sharing upon request after the acceptance of the manuscript

## ACKNOWLEDGMENTS

HCWs from Laiko General Hospital and Onassis Cardiac Surgery Center

P. Minogiannis, I. Papaparaskevas, I. Boletis - Onassis Cardiac Surgery Center

Z. Moschidis, E. Kokolesis, M. Katsimicha – Hellenic Scientific Society for the Study of AIDS, Sexually Transmitted and Emerging Diseases

## FUNDING

The study was funded by:

Onassis Cardiac Surgery Center

Union of Greek Ship Owners

SB Bioanalytica S.A.

Hellenic Scientific Society for the Study of AIDS, Sexually Transmitted and Emerging Diseases

---

**Supplementary Table 1.** Median (25^th^-75^th^) levels of anti-SARS-CoV-2 RBD IgG by time (days) from 2^nd^ dose of BNT162b2 vaccine

|  | N (%) | Median (25th-75th) (AU/ml) | p<sup>1</sup> |
| --- | --- | --- | --- |
| **Days from second dose** |  |  | **0.007** |
| 5-7 | 131 (15.0) | 15,107 (6,736 - 26,119) |  |
| 8 | 142 (16.3) | 16,503 (9,678 - 29,683) |  |
| 9 | 128 (14.7) | 17,514 (9,867 - 27,522) |  |
| 10 | 107 (12.3) | 17,081 (9,531 - 28,318) |  |
| 11 | 56 (6.4) | 19,830 (13,525 - 30,796) |  |
| 12 | 88 (10.1) | 15,229 (10,274 - 29,429) |  |
| 13 | 80 (9.2) | 15,706 (8,557 - 23,497) |  |
| 14 | 92 (10.6) | 12,462 (7,519 - 27,151) |  |
| 15-17 | 47 (5.4) | 10,330 (5,529 - 18,880) |  |

*Abbreviations: 25th–75th, 25th and 75th percentiles; AU/ml, arbitrary units per milliliter*
<sup>1</sup>*Nonparametric Kruskal-Wallis test*

## Figure Legends

**Supplementary Figure 1A.**
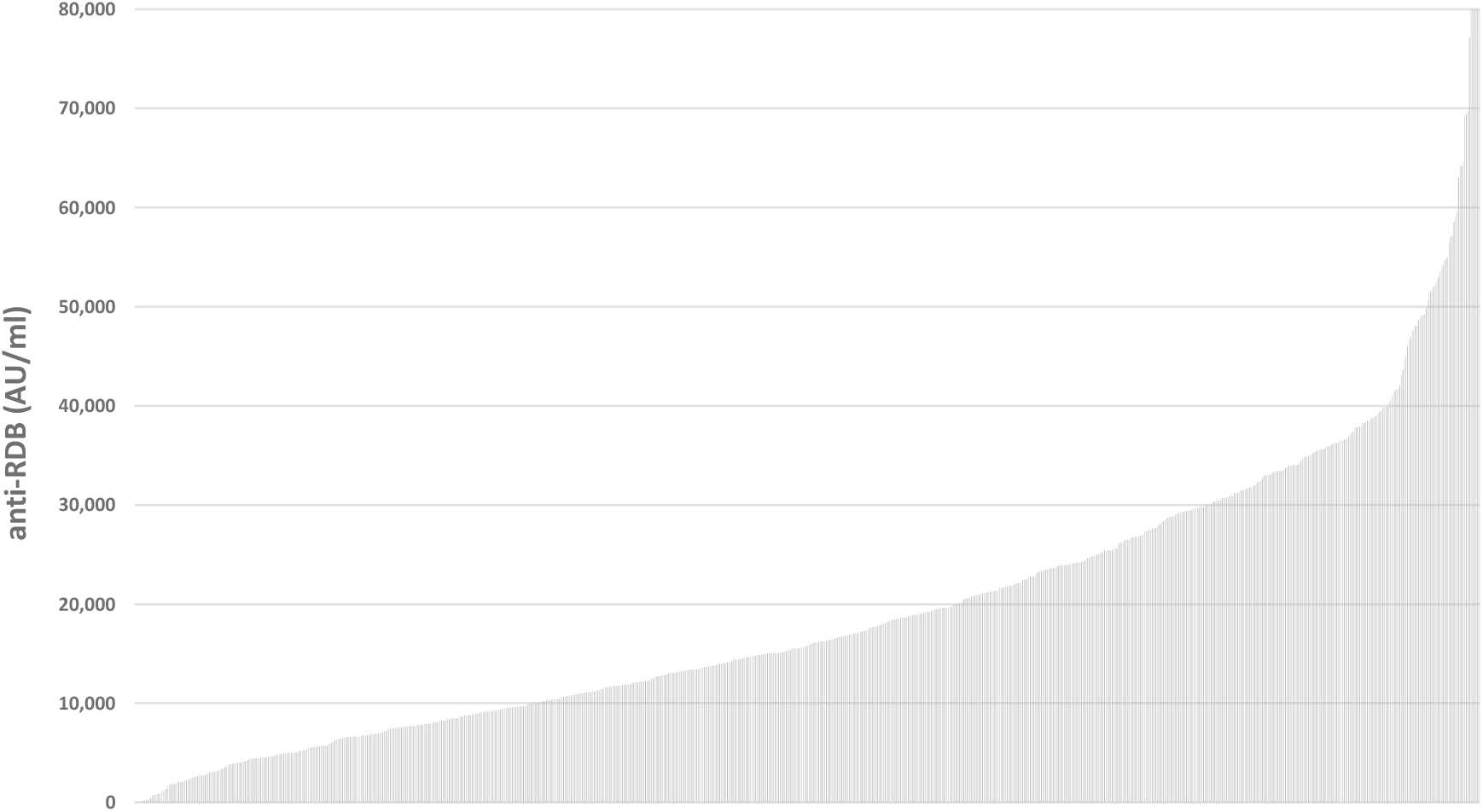
Cumulative distribution of anti-SARS-CoV-2 RBD AU/ml in vaccinated health care workers 5-17 days after the 2nd dose of BNT162b2.

**Supplementary Figure 1B.**
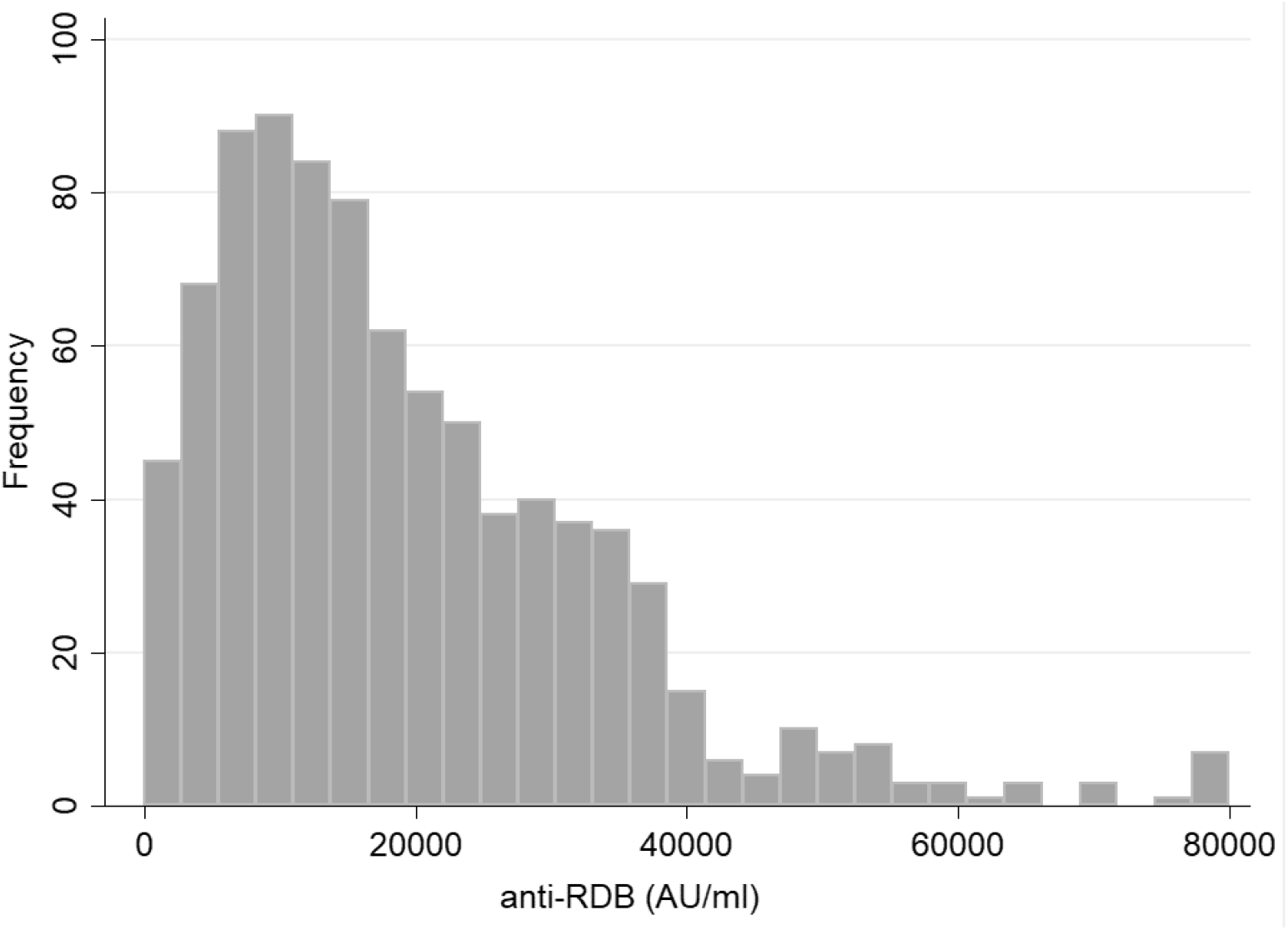
Frequency distribution of anti-SARS-CoV-2 RBD AU/ml in vaccinated health care workers 5-17 days after the 2nd dose of BNT162b2.

